# HLA-A*03:01 is associated with increased risk of fever, chills, and more severe reaction to Pfizer-BioNTech COVID-19 vaccination

**DOI:** 10.1101/2021.11.16.21266408

**Authors:** Alexandre Bolze, Iva Neveux, Kelly M. Schiabor Barrett, Simon White, Magnus Isaksson, Shaun Dabe, William Lee, Joseph J. Grzymski, Nicole L. Washington, Elizabeth T. Cirulli

## Abstract

COVID-19 vaccines are safe and highly effective, but some individuals experience unpleasant reactions to vaccination. As the majority of adults in the US have received a COVID-19 vaccine this year, there is an unprecedented opportunity to study the genetics of reactions to vaccination via surveys of individuals who are already part of genetic research studies. Here, we have queried 17,440 participants in the Helix DNA Discovery Project and Healthy Nevada Project about their reactions to COVID-19 vaccination. Our GWAS identifies an association between severe difficulties with daily routine after vaccination and HLA-A*03:01. This association was statistically significant only for those who received the Pfizer-BioNTech vaccine (BNT162b2; p=4.70E-11), but showed a trending association in those who received the Moderna vaccine (mRNA-1273; p=0.005) despite similar sample sizes for study. In Pfizer-BioNTech recipients, HLA-A*03:01 was associated with a two-fold increase in risk of severe vaccine reactions. The effect was consistent across ages, sexes, and whether the person had previously had a COVID-19 infection. The reactions experienced by HLA-A*03:01 carriers were driven by associations with chills, fever, fatigue, and in general feeling unwell.

## Introduction

Less than one year after the first publication of a SARS-CoV-2 sequence, COVID-19 vaccines were developed, clinically tested and authorized to be administered in the general population. Within months, hundreds of millions of adults worldwide were vaccinated, and rates of hospitalization among vaccinated individuals dropped precipitously; potential side effects were mild^1^. During the clinical trials of mRNA vaccines (Pfizer-BioNTech BNT162b2 and Moderna mRNA-1273), both local reactions, such as pain at the injection site, and systemic symptoms such as fatigue, fever, chills, and myalgia were observed in some participants ^2, 3^. Both clinical trials showed that only a small fraction of these reactions could be categorized as severe, and most of the severe reactions followed the second dose. These studies raise the question of what factors explain the interindividual variability of reactions following COVID-19 vaccination.

Younger age and a personal history of SARS-CoV-2 infection prior to vaccination are two factors leading to increased reactogenicity to the vaccine^4^. However, these factors alone do not explain the large interindividual variability in the degree of severity of the reaction following COVID-19 vaccination. The aim of this study is therefore to identify factors associated with severe reactions following COVID-19 vaccination. Similarly to our hypothesis that genetic factors play a role in the severity of COVID-19 disease^5, 6^, we hypothesized that genetic factors help explain differences in reactions following COVID-19 vaccination. To test our hypothesis, we administered online surveys to 17,440 participants from the Helix DNA Discovery project and the Healthy Nevada Project asking about their vaccination status and their reactions following the vaccine. These surveys included questions about 4 local and 20 systemic symptoms, as well as the overall severity of the reaction and its impact on daily activities in the days following vaccination. These participants were previously sequenced with Helix Exome+ assay, which allowed us to perform rare and common variant genetic associations^7^.

## Methods

### Cohort and survey

We administered an online survey in June and September of 2021 and received responses from 8,125 Helix DNA Discovery Project participants and 9,315 Healthy Nevada Project participants (Table 1)^8, 9^. These are unselected Helix customers and patients in the Renown Health System who chose to consent to participate in research projects and respond to our survey. The survey takes approximately 10 minutes to complete and can be found in the supplement. The participants in this cohort are aged 18 to 89+, 65% are female, and 85% are of European genetic ancestry (Table 1). The Helix DNA Discovery Project study was reviewed and approved by Western Institutional Review Board. The HNP study was reviewed and approved by the University of Nevada, Reno Institutional Review Board.

**Table 1.** Study and cohort information.

|  |  |
| --- | --- |
| Sample size | 17,440 |
| Median age (range) | 58 (18-89+) |
| N female (%*) | 10,402 (64.9%) |
| Genetic ancestry N (%*) |  |
| African | 264 (1.6%) |
| East Asian | 301 (1.9%) |
| European | 13,643 (84.9%) |
| Hispanic | 1,391 (8.7%) |
| South Asian | 60 (0.4%) |
| Other / mixed ancestry | 403 (2.5%) |
| N with COVID-19 vaccination (%) |  |
| Pfizer | 8,041 (46.1%) |
| Moderna | 7,086 (40.6%) |
| J&J | 790 (4.5%) |
| Other/Unsure | 227 (1.3%) |
| N with positive COVID-19 test before vaccination (%) | 1,280 (7.3%) |
| N reporting vaccine reactions (%) |  |
| 4- Extreme difficulties / unable to perform daily routine | 1,029 (8.0%) |
| 3- Severe difficulties with daily routine | 1,237 (9.6%) |
| 2- Moderate difficulties with daily routine | 2,597 (20.2%) |
| 1- Mild difficulties with daily routine | 3,502 (27.2%) |
| 0- No difficulties with daily routine | 4,500 (35.0%) |

\*The total here is adjusted to remove individuals who do not have their sex and ethnicity available: this demographic information was collected separately and was not yet available for some individuals for this round of analysis.

Respondents indicated whether they had been vaccinated and which vaccine type they had received. They rated the severity of their vaccine response as indicated in Table 1. They also answered questions about 24 specific side-effects that can occur after COVID-19 vaccination (see Supplemental survey). There were 3,323 individuals who took the survey in both June and September, updating their COVID-19 vaccination status and infection status. For these individuals, the highest severity score and all reactions were used as a phenotype, regardless of in which survey either were reported.

### Genotyping

DNA samples were sequenced and analyzed at Helix using the Exome+^®^ assay as previously described^10^. Imputation of common variants was performed by pre-phasing samples and then imputing. Pre-phasing was performed using reference databases, which include the 1000 Genomes Phase 3 data. This was followed by genotype imputation for all 1000 Genomes Phase 3 sites with MAF>=1% that have genotype quality (GQ) values less than 20. Imputation results were then filtered for quality (GP>=0.95) so that only high precision imputed variant calls were reported^7^.

HLA genotypes for A, B, C, DPB1, DQA1, DQB1, and DRB1 were imputed using HIBAG with the default recommendations^11^. Individual genotypes were imputed using the model for the appropriate genetic ancestry for each individual^11^. Probabilities higher than 0.5 were used as genotype calls.

### Genetic analysis

We used Regenie for the genetic analysis^12^. Briefly, this method builds a whole genome regression model using common variants to account for the effects of relatedness and population stratification, and it accounts for situations where there is an extreme case-control imbalance, which can lead to test statistic inflation with other analysis methods. The covariates we included were age, sex, age*sex, age*age, sex*age*age, 10 PCs, type of COVID-19 vaccine, whether the individual had COVID-19 prior to getting vaccinated, and bioinformatics pipeline version. We only utilized phenotypes with at least 50 cases, with a minimum minor allele count (MAC) cutoff of 5.

As previously described, a representative set of 184,445 coding and noncoding LD-pruned, high-quality common variants were identified for building PCs and the whole genome regression model^10^. Each genetic ancestry group was analyzed separately: African, East Asian, European, Hispanic, South Asian.

## Results

### A minority of individuals have severe difficulties with daily routine following COVID-19 vaccination

We received survey responses from 8,125 Helix participants and 9,315 HNP participants in June and September 2021. As a control, we first checked whether we observed the same trends as reported in the clinical trials of the Pfizer-BioNTech, Moderna and J&J COVID-19 vaccines. The large majority of participants in these clinical trials did not have a personal history of SARS-CoV-2 infection. To assess the frequency of reactions in our cohort, we therefore restricted our initial analysis to participants who reported no previous infection by SARS-CoV-2. Our results were in agreement with what was previously reported^2, 3, 13^. For the Pfizer-BioNTech vaccine, we observed that only 2.7% of individuals (152 of 5,590) had severe or extreme difficulties with their daily routine after the first dose, and 10.2% (569 of 5,583) had severe or extreme difficulties after the second dose (Figure S1). For the Moderna vaccine, we observed that 4.4% of individuals (243 of 5,575) had severe or extreme difficulties after the first dose, and this number grew to 19.5% (1,083 of 5,558) after the second dose (Figure S1). For the J&J vaccine, we observed that 17.7% (80 of 452) had severe difficulties after the single dose (Figure S1). Moreover, we observed a higher number of severe reactions in respondents 18-55 years old compared to those over 55 (Figure S1). These analyses supported the observation that there are large differences in reactions to COVID-19 vaccination between individuals, and validated our self-reported survey as a reliable tool to investigate the genetic basis of these differences.

### GWAS identifies HLA region associated with severe reaction to COVID-19 vaccination

To test our hypothesis that genetic variation drives some of the differences seen in vaccine reaction, we performed a genome-wide association study (GWAS) using a phenotype of extreme or severe difficulties with daily routine (1,709 cases) compared to no or mild difficulties (6,203 controls). Individuals reporting moderate reaction were excluded from the GWAS (Table 1). Our analysis of 12,602,603 SNPs identified 188 genome-wide significant variants on chromosome 6 (p<5e-8; Figure 1). The lead SNP was rs144943243 / chr6:29820015:AAAAT:A, p=3.51E-11. This variant is in a region of AAAT repeats found upstream of *HLA-G*, with a MAF of 24% in the European genetic ancestry subset of our cohort.

**Figure 1.**
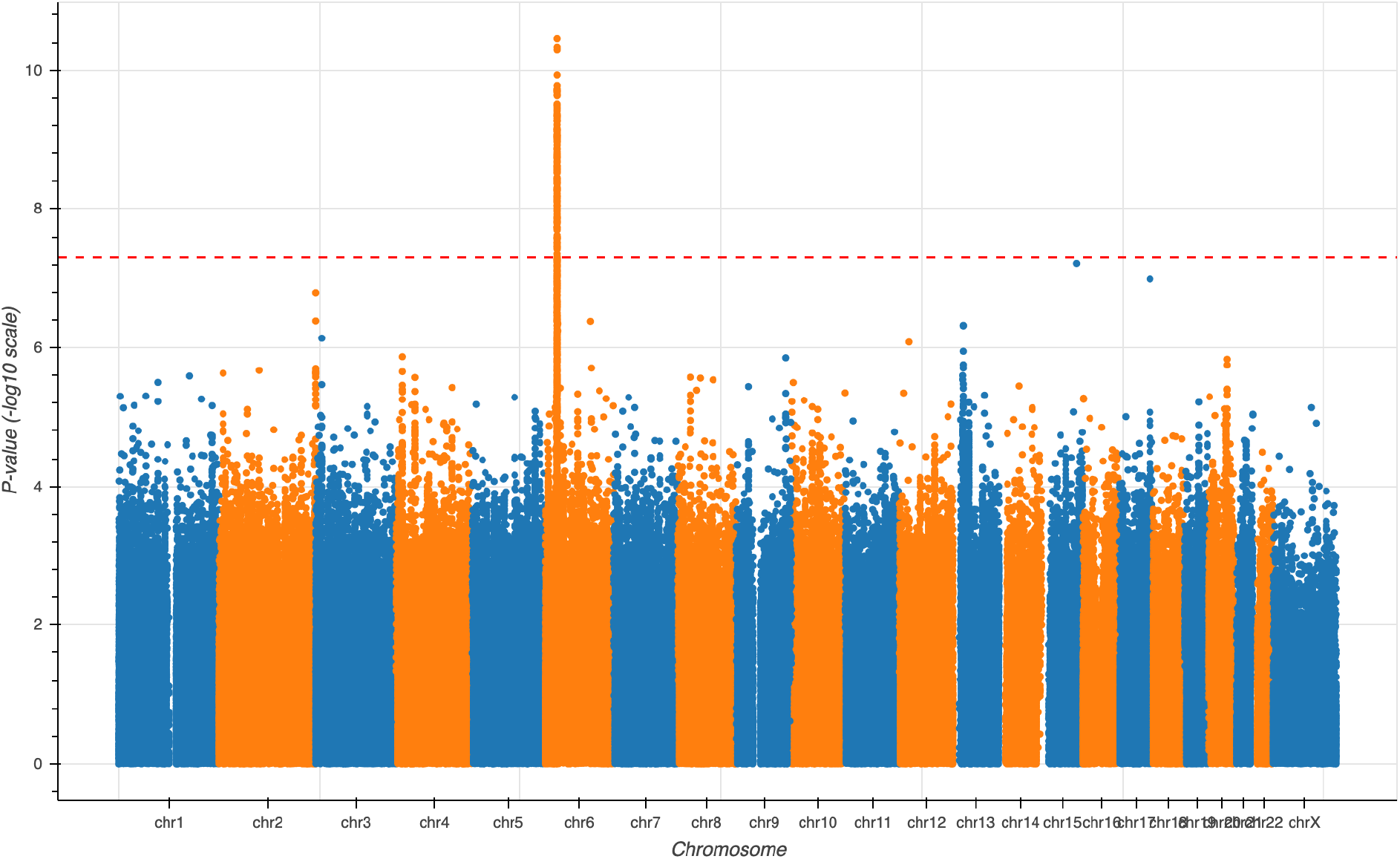
Manhattan plot for main phenotype of severe / extreme vaccine reaction against mild or no reaction to any vaccination event with Pfizer-BioNTech, Moderna, J&J, or other COVID-19 vaccines. The lambda GC was 1.07.

We next analyzed imputed HLA types against the phenotype to identify whether a specific HLA type was associated with the GWAS signal. We identified a significant association with HLA-A*03:01 (p=5.00E-11), which had a MAF of 15% in the European genetic ancestry subset of our cohort. The distribution of HLA-A*03:01 across the globe is shown in Figure S2^14^. A regression including both this HLA type and rs144943243 identified similar signals for each variant, with rs144943243 retaining a better p-value in a joint analysis. The subsequent analyses assessing the impact of the genetic variant by phenotype lead to similar results for rs144943243 and HLA-A*03:01. In this manuscript, we decided to show the results for HLA-A*03:01, a common allele for the well-studied *HLA-A* gene

### HLA-A*03:01 association is specific to vaccine type

HLA-A*03:01 had an odds ratio (OR) of 1.5 for individuals to experience severe / extreme difficulties with daily routine after vaccination as opposed to a mild or no difficulties. We found that this effect was additive, with individuals with 1 copy of HLA-A*03:01 having a phenotype intermediate to those with 0 or 2 copies (Figure 2). We found the effect of this variant to track similarly across age, sex, and whether the person had a history of SARS-CoV-2 infection prior to vaccination (Figure 2, S3). However, our analyses beyond European (p=5.00E-11, OR=1.6) and Hispanic (p=2.93E-04, OR=2.7) genetic ancestries were underpowered to identify associations (Table S1). We also observed that most of the effect seems to occur at the second dose for two-dose vaccines (Figure S4), and that severe reactions subsided within 2 days for 66% of those who experienced severe reactions (Figure S5).

**Figure 2.**
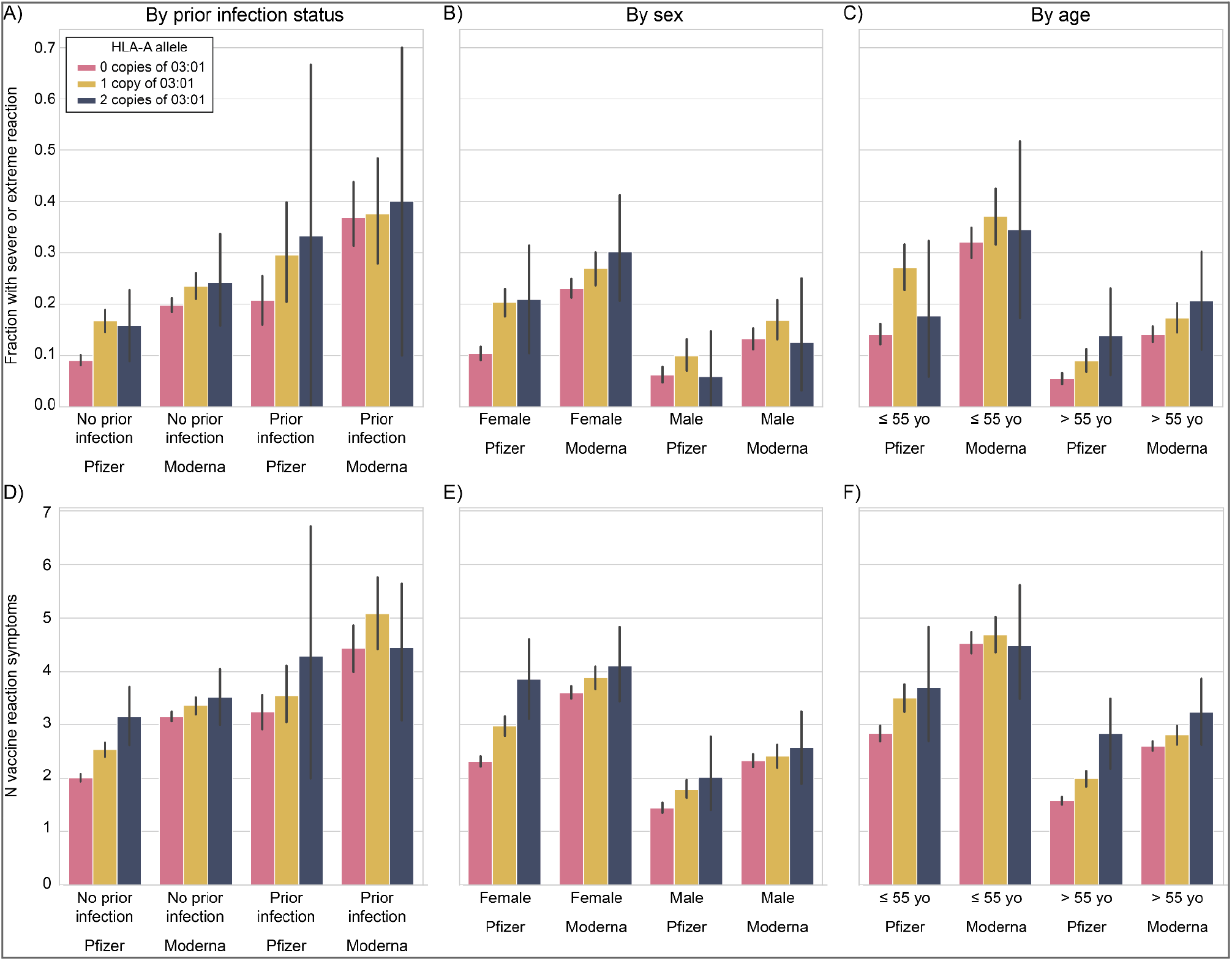
Risk of vaccine side effects by HLA-A*03:01 genotype and vaccine type. A and D) Broken down by whether they had COVID-19 prior to vaccination. B and E) Broken down by sex (COVID-19 prior to vaccine excluded). C and F) By age (COVID-19 prior to vaccine excluded). Top row (A-C): Fraction with severe or extreme reaction; Bottom row (D-F): Number of vaccine reaction symptoms per person. European genetic ancestry with Pfizer-BioNTech or Moderna only shown.

Importantly, we identified that the association signal came almost entirely from reactions to the Pfizer-BioNTech vaccine (HLA-A*03:01 p=4.70E-11, OR=2.07; best Pfizer-BioNTech SNP chr6:29945053:T:C / rs2571381 p=1.16E-12), with very little impact on Moderna vaccine response (p=0.005, OR=1.32) despite similar sample sizes for analysis (Pfizer 552 cases vs. 3,142 controls; Moderna 1,018 cases vs. 2,592 controls). While the main phenotype of severe / extreme difficulties with daily routine after vaccination was more common in Moderna recipients compared to Pfizer-BioNTech recipients, even restricting to a more stringent case definition did not result in statistical significance for the Moderna subset (p=0.008, OR=1.5; 473 extreme reaction cases vs. 1,425 no reaction controls). The sample sizes for those who received J&J or other vaccinations were too low to clearly assess an association. Detailed counts of reaction severity split by genetic ancestry, HLA-A*03:01 status, and vaccine type are reported in Table S1.

### Specific vaccine reaction phenotypes are associated with HLA-A*03:01

To understand the severity scores more deeply, we next analyzed the individual symptoms that participants reported as occurring after receiving the vaccine, split into Pfizer-BioNTech and Moderna subsets. We identified that in Pfizer-BioNTech recipients, HLA-A*03:01 was most strongly associated with increased risk of fever (p=1.76E-14), chills (p=6.15E-13), feeling unwell (p=5.73E-10) and fatigue (p=5.60E-07) after receiving the vaccine (Figure 3). The ORs for these reactions ranged from 1.44-2.09, with 41% of those with 1 HLA-A*03:01 copy who received a Pfizer vaccine having at least two of these four symptoms, 27% of those with 2 copies, and 19% of those with 0 copies. Associations with joint pain, headache, swollen lymph nodes, headache and nausea were less predictive but still statistically significant (p<0.001). In contrast, none of these symptoms were significantly associated with HLA-A*03:01 in Moderna recipients (Figure 3).

**Figure 3.**
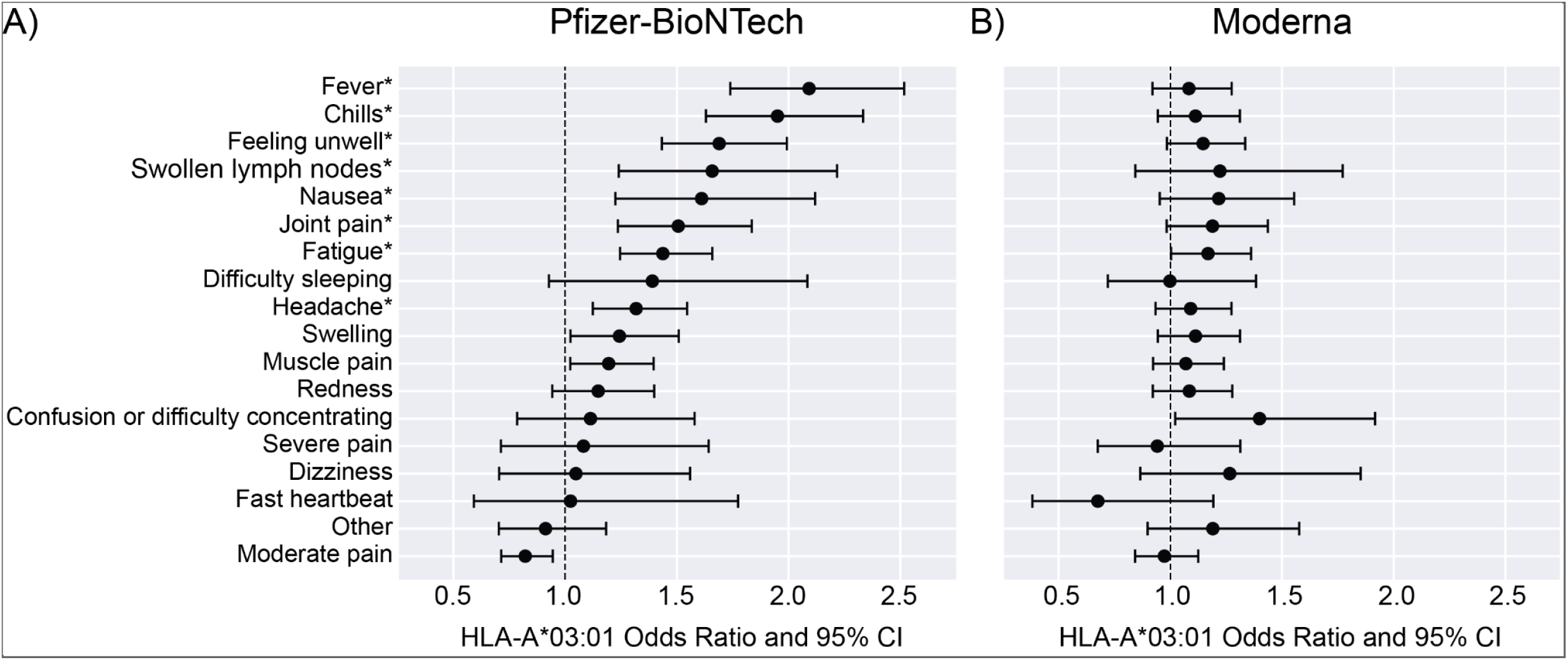
Odds ratios and 95% Confidence Interval (CI) for specific vaccine responses in an additive genetic analysis (regenie) of HLA-A*03:01 in European ancestry individuals (n=9,636). *p<0.001 in Pfizer-BioNTech recipients (A); no associations were significant in Moderna recipients (B).

## Discussion

Here, we identified an HLA type, HLA-A*03:01, with a strong association with reactions to COVID-19 vaccines. We find that, all else being equal, individuals with this HLA type who received the Pfizer-BioNTech vaccine are approximately twice as likely to have severe or extreme difficulties with their daily routine following COVID-19 vaccination. Chills and fever were the two specific side effects that were most enriched in individuals carrying 1 or 2 copies of HLA-A*03:01 compared to individuals with two other HLA-A alleles. This association was present in the Helix DNA Discovery cohort, as well as the Healthy Nevada Project cohort. We find this association to trend across age groups, sex, and whether the person had a personal history of COVID-19 prior to vaccination, all of which are known to be associated with severity of vaccine reaction. We find that the effect of this variant was almost entirely driven by participants who had received the Pfizer-BioNTech vaccine. The signal for the association was very weak in those receiving the Moderna vaccine and did not approach genome-wide significance, despite a sample size roughly equal to that of the Pfizer vaccine recipients. Our sample size for J&J and other vaccines were too low for adequate analysis power.

The difference of effect size and significance between the Pfizer-BioNTech and Moderna vaccines was surprising given the similarities between the two mRNA vaccines. It is possible that the higher dose of the Moderna vaccine leads to a stronger innate immune activation, which would be independent of the role of the HLA-A*03:01 allele, which is more likely to be involved in the adaptive immune response. This stronger innate immune activation may dilute the effect of HLA-A*03:01 in recipients of the Moderna vaccine. It could also be that the increased prevalence of vaccine reactions in Moderna recipients make the signal harder to identify in that subgroup (Figure 2, S1, S4). Replication of the result in other cohorts, with results broken down by COVID-19 vaccine types will be informative to hypothesize further about potential mechanisms leading to these severe reactions. An effect, even of different intensity, in all types of COVID-19 vaccines could be linked to a common peptide derived from all vaccines showing an association. Importantly, as we were preparing this manuscript, 23andMe published on their blog (https://blog.23andme.com/23andme-research/reaction-to-covid-vaccine/, accessed 14 November 2021) showing that HLA-A*03:01 was also the strongest genetic association with COVID-19 vaccine response in their cohort with a p-value p=1.5E-130. No additional details regarding the effect by vaccine type, dose, or symptoms were available yet.

HLA-A*03:01 is the third most frequent HLA-A allele in the population^14^. HLA-A*03:01 was reported to be associated with haemochromatosis in the UK Biobank cohort^15^. However, this association could be due to the fact that HLA-A3/B7 is the ancestral haplotype from which the *HFE* p.C282Y pathogenic variant originated^16^. Neither HLA-A*03:01 nor the other top hits from our GWAS, such as rs2571381, have been associated with COVID-19 severity or susceptibility to infection^5^. This allele was also not reported to be associated with non-COVID vaccine reactions, or reactions to medications based on our literature search.

Associations of class I HLA alleles and reactions to medications have been reported in the past. For example, HLA-B*57:01 is linked to hypersensitivity to HIV-1 reverse-transcriptase inhibitor abacavir^17, 18^. However, to our knowledge, very few studies have been published reporting genetic associations with reactogenicity after non live-attenuated vaccines. In the past, a few studies have investigated the genetics of response to vaccines by looking at the levels of antibodies after a certain time period in vaccine recipients, often identifying associations with HLA genes^19, 20^. Other studies have elucidated why some patients presented with a rare life-threatening disease following vaccinations with live-attenuated vaccines^21^. For example, genetic defects in the IL-12 dependent IFN-gamma pathway cause BCGitis after BCG vaccination^22^. The difficulty of collecting appropriate phenotype information on the reaction to a vaccine with standard medical data or electronic health records could be one reason to explain the small number of these studies. Another reason may be the difficulty to justify sequencing individuals to investigate a transient phenotype. The ability for us to ask new questions and gain new biological insights highlight the importance of continuing to survey and engage participants who are enrolled in ongoing genetic research projects. The closer we get to a state where a recipient’s genetic profile is readily available within a health system’s records, serving up information about these types of associations could be highly valuable in educating and preparing anxious vaccine recipients about what to expect. Additionally, the more we understand about these genetic associations, the more vaccine manufacturers can incorporate and attempt to mitigate these potential reactions into their vaccine development process.

Lastly, it is important to emphasize that our study defined a severe reaction to the vaccine as a reaction that would interfere with daily routine shortly after receiving the vaccine. These reactions were symptoms such as chills or fever, which cannot be compared with the severity of COVID-19 disease experienced by many individuals. COVID-19 vaccines have consistently been shown to be safe and very effective to prevent hospitalizations and life-threatening disease following SARS-CoV-2 infections^2, 3, 13, 23^.

## Data Availability

All data produced in the present study are available upon reasonable request to the authors

## Acknowledgements

Funding was provided to DRI by the Nevada Governor’s Office of Economic Development. Funding was provided to the Renown Institute for Health Innovation by Renown Health and the Renown Health Foundation. We acknowledge the entire Helix Bioinformatics team for their contributions to the production exome sequencing pipeline. We thank C. Clinton, KT Farley and E. Levin for their contribution running the Helix DNA Discovery Project and the work with the IRB. We thank all of the genomic representatives of the Healthy Nevada Project (HNP). We thank Renown Health and DRI marketing for helping to launch the HNP. We thank J. Fellay for comments on the manuscript.

## Supplementary Information

**Figure S1.**
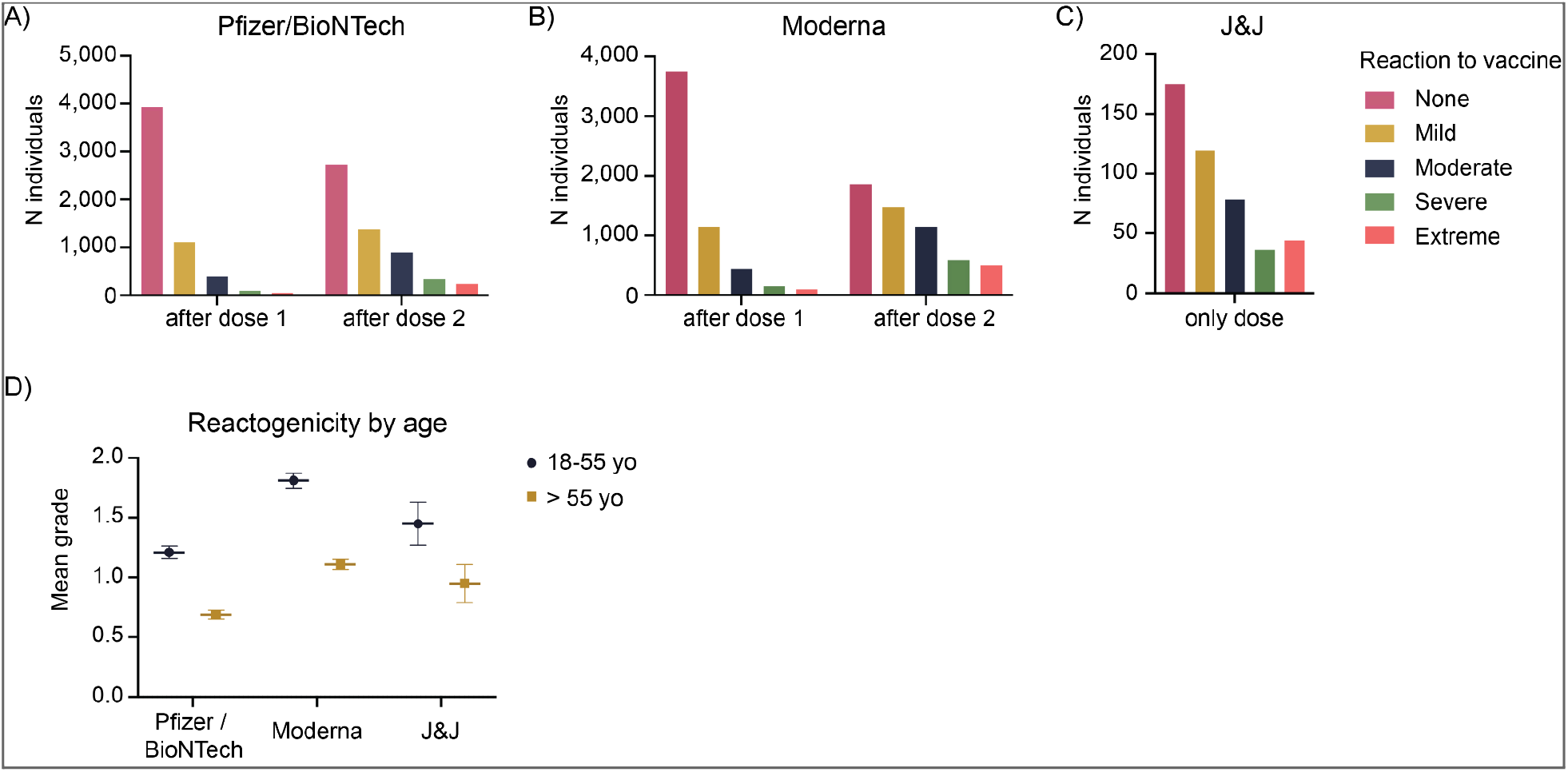
Self-reported severity of vaccine reaction by type of vaccine, and by age. This analysis was limited to participants without a personal history of SARS-CoV-2 infection prior to vaccination. **A-C)** Reaction severity after dose 1 (left) and after dose 2 (right) of Pfizer-BioNTech vaccine (A); after dose 1 (left) and after dose 2 (right) of Moderna vaccine (B); after J&J vaccine (C). Columns of different colors represent the number of individuals who self-reported with a reaction of a given severity. Purple: No difficulties with daily routine. Blue: Mild difficulties with daily routine. Green: Moderate difficulties with daily routine. Orange: Severe difficulties with daily routine. Red: Extreme difficulties / unable to perform daily routine. **D)** Reactogenicity by age. For each vaccine, the mean self-reported reaction grade is plotted. 0.0: no reaction; 1.0: mild reaction; 2.0: moderate reaction; 3.0: severe reaction; 4.0 extreme reaction. In blue (rounds) for individuals 18 to 55 years old, and in orange (squares) for individuals over 55 years old. Left panel: Pfizer-BioNTech vaccine, Middle panel: Moderna vaccine. Right panel: J&J vaccine.

**Figure S2.**
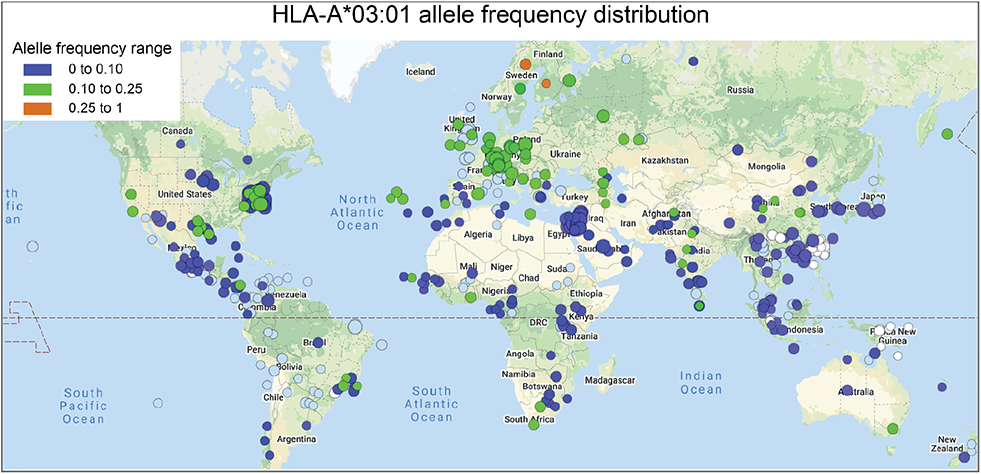
Distribution of HLA-A*03:01 across the globe. Downloaded from http://www.allelefrequencies.net/hla6008a.asp?hla_allele=A*03:01 on November 15, 2021.

**Figure S3.**
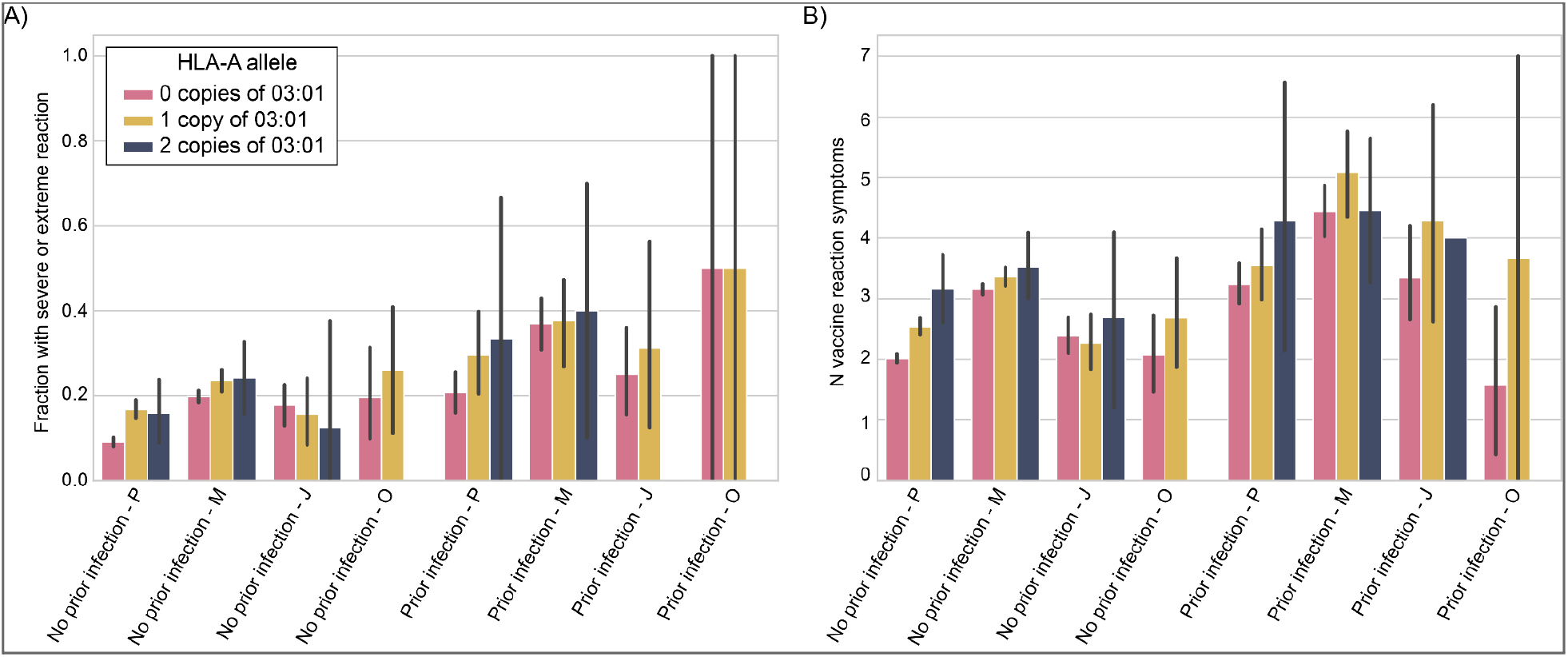
Rate of severe / extreme vaccine reaction by vaccine manufacturer, HLA-A*03:01 genotype, and whether participants had COVID-19 prior to the vaccine. P=Pfizer-BioNTech, M=Moderna, J=Johnson&Johnson, O=Other/Unsure. Plot is restricted to European genetic ancestry individuals.

**Figure S4.**
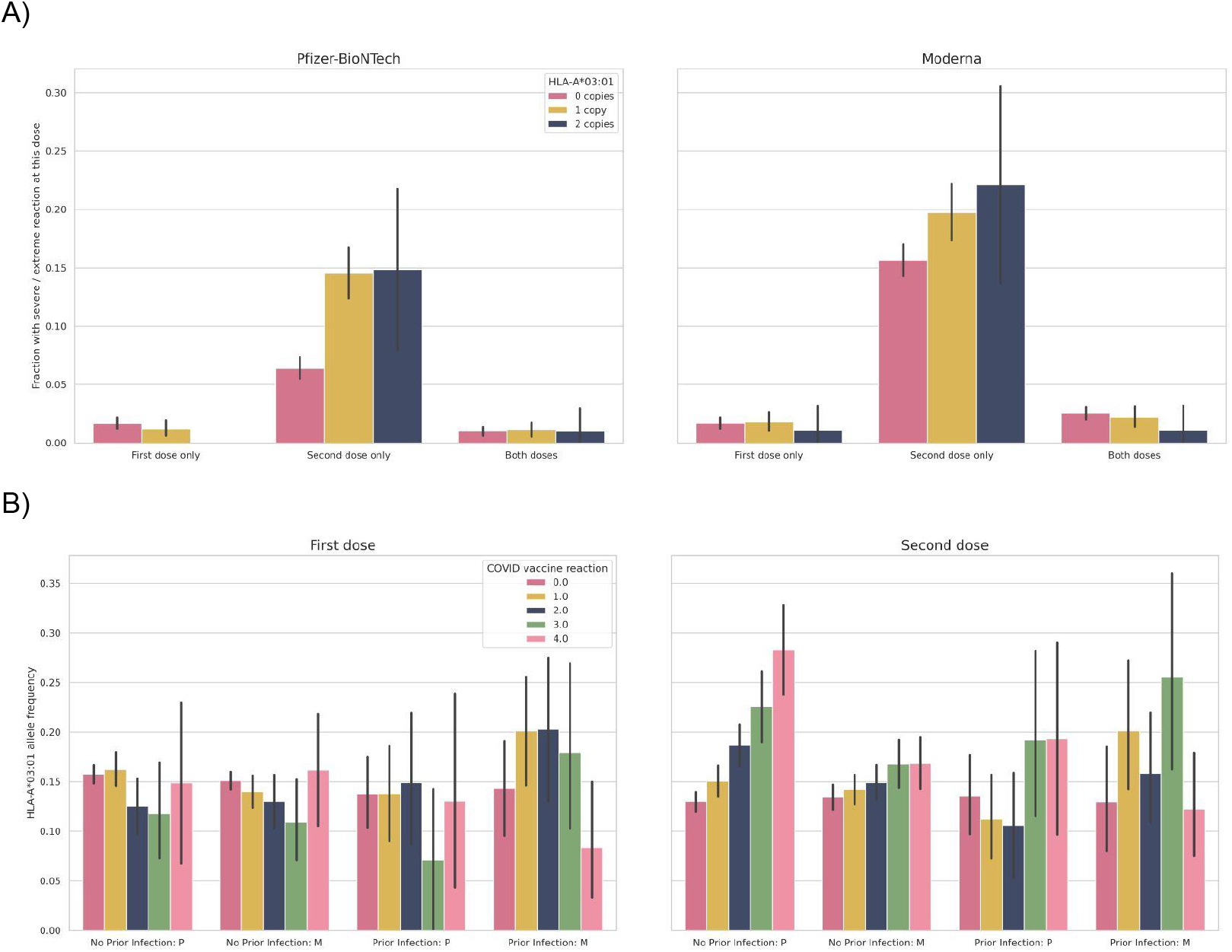
Reaction by first and second dose. A) Percent of vaccine recipients experiencing severe / extreme vaccine reaction at only the 1st dose, only the 2nd dose, or at both doses. B) MAF of HLA-A*03:01 for individuals who rated their vaccine reaction as none (0), mild (1), moderate (2), severe (3) or extreme (4) according to Table 1). At the second dose, individuals who received Pfizer had the clearest association with HLA-A*03:01. Plot is restricted to European ancestry individuals.

**Figure S5.**
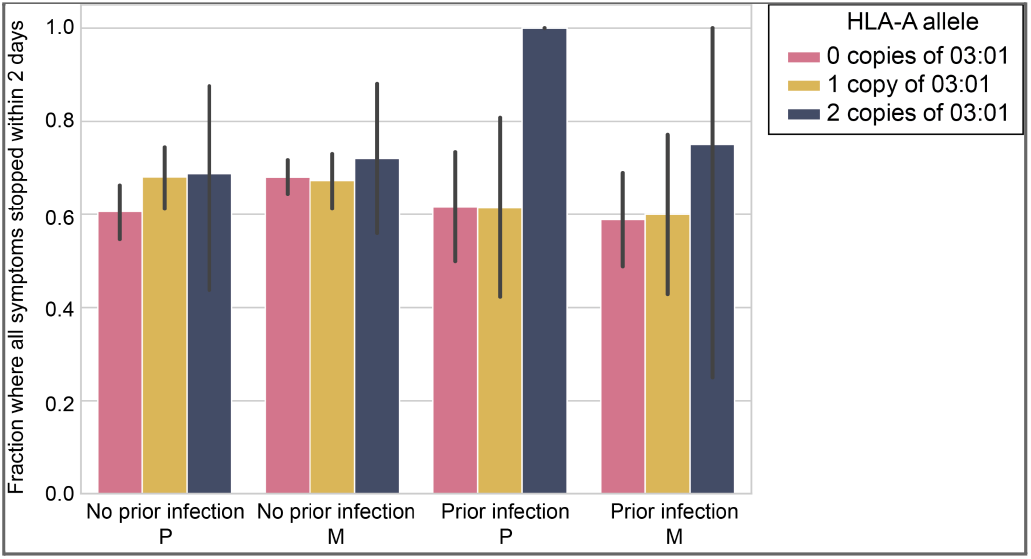
Fraction of individuals with severe / extreme vaccine reactions whose severe reactions subsided within two days. Individuals are split by which vaccine they received. Plot is restricted to European ancestry individuals.

**Table S1:** Counts of individuals by maximum reaction severity and grouped by genetic ancestry

## Survey text

**“Thank you for taking time on these questions. Your answers will help Helix researchers and the healthcare community learn more about COVID-19. This should only take about 15 minutes. To protect your privacy, your answers will be kept nameless.”**

Have you received a COVID-19 vaccine?

Yes / No

**(this section only if Yes; otherwise skip to 1 in next section)**

1a. Which vaccine did you receive?

Pfizer-BioNTech (2 doses)
Moderna (2 doses)
Johnson & Johnson (1 dose)
Other (please specify)
Unsure

(if 1 dose) You are considered fully vaccinated 2 weeks after your vaccine. Has it been at least 2 weeks since your vaccine?

Yes / No

(if 2 dose) You are considered fully vaccinated 2 weeks after your second vaccine. Has it been at least 2 weeks since your second vaccine?

Yes / No

**(this section only if Yes; otherwise skip to 1 in next section)**

1b. Did you take any of the following painkillers immediately before or within one day after receiving the vaccine?

Ibuprofen (including Advil, Motrin)

Naproxen (including Aleve)

Aspirin

Acetaminophen (including Tylenol)

COX inhibitors (including Celebrex)

Cannabis (including marijuana)

Other (please specify)

1. Did you experience any of the following symptoms after receiving the vaccine [check all that apply]:

Swelling at the injection site
Redness or discoloration at the injection site
Severe pain at the injection site
Moderate pain at the injection site
Muscle pain
Joint pain
Fatigue
Fever
Headache
Confusion or difficulty concentrating
Chills
Nausea
Vomiting
Intense hunger
Swollen lymph nodes / glands
Feeling unwell
Difficulty breathing
Swelling of your face or throat
A fast heartbeat
A bad rash all over your body
Dizziness
Difficulty sleeping
Delayed menstrual cycle
Early menstrual cycle
Other (please specify)
None
2. When at their worst, how much did the symptoms that you experienced after the COVID-19 vaccine interfere with your daily routine? (for 1 dose vaccine only)

Extreme difficulties / unable to perform daily routine
Severe difficulties with daily routine
Moderate difficulties with daily routine
Mild difficulties with daily routine
No difficulties with daily routine
3. When at their worst, how much did the symptoms that you experienced after the first dose of the COVID-19 vaccine interfere with your daily routine? (for 2 dose vaccine only)

Extreme difficulties / unable to perform daily routine
Severe difficulties with daily routine
Moderate difficulties with daily routine
Mild difficulties with daily routine
No difficulties with daily routine
4. When at their worst, how much did the symptoms that you experienced after the second dose of the COVID-19 vaccine interfere with your daily routine? (for 2 dose vaccine only)

Extreme difficulties / unable to perform daily routine
Severe difficulties with daily routine
Moderate difficulties with daily routine
Mild difficulties with daily routine
No difficulties with daily routine

(for 2 dose vaccine only, state:) For the following questions, please respond about the worst symptoms you experienced, whether after dose 1 or 2.

6a How long after receiving the COVID-19 vaccine did these symptoms begin?

Within 4 hours

Within 3 days

After more than 3 days

6b Once they started, how long did it take for the MOST SEVERE symptoms of the COVID-19 vaccine to go away?

Less than two days
Two to seven days
More than a week

6c Once they started, how long did it take for ALL of the symptoms of the COVID-19 vaccine to go away?

Less than two days
Two to seven days
More than a week

5. Have you ever been sick with COVID-19?

Yes / No

5a if Yes to 5: Have you had COVID-19 more than once?

Yes / No

5b if Yes to 5: Did you have COVID-19 before or after being fully vaccinated?

Before / after

6 if no to 5: At any point since being fully vaccinated, have you had a new onset of an illness?

Yes
No
7 (if yes to 5a) Which of the following best describes your COVID-19 history:

I had COVID-19 twice, both times before being fully vaccinated
I had COVID-19 twice, both times after being fully vaccinated
I had COVID-19 twice, once before and once after being fully vaccinated
Other (specify)
8 if had COVID before vaccination: Did you have ongoing symptoms from COVID-19 in the week before you were given your first COVID-19 vaccine injection?

Yes / No 8a. If yes What type of ongoing COVID-19 symptoms did you have in the week before you were given the COVID-19 vaccine injection? Acute symptoms from an early phase of the illness, such as fever and cough Long-term symptoms from a lingering illness, such as fatigue or confusion Other (please specify): 8a if Yes Please tell us whether your symptoms from your COVID-19 illness changed 2 weeks or later after having your first COVID-19 vaccine injection. Yes – They all got better Yes – Some of them got better No change Yes – Some of them got worse Yes – They all got worse Some improved and others got worse if had COVID, skip to 1.1 below If had had non-COVID illness, skip to 1.2 If had no illness, skip to 1.1b.

**(end of Yes to vaccine section)**

1 Have you ever been sick with COVID-19?

Answers
Yes, I have received a positive COVID-19 test
Yes, although I have not received a positive COVID-19 test
No, although I think that I was exposed to COVID-19
No, and I do not think that I was exposed to COVID-19
Unsure
1a (if positive test) Have you had COVID-19 more than once?

Yes
No
1.1 (if exposed) How were you exposed to COVID-19? Please check all boxes that apply. Series of Check boxes:

_I’ve been in close physical contact with someone while they had coronavirus
_I’ve been in the same room or vehicle as someone while they had coronavirus
_I’ve lived with someone while they had coronavirus
_I was informed through contact tracing that I was exposed
_I am a healthcare worker who works with coronavirus patients
_No known exposure
_Other (please specify)
1.1a (if positive) When at their worst, how much did your COVID-19 symptoms interfere with your daily routine?

Extreme difficulties / unable to perform daily routine
Severe difficulties with daily routine
Moderate difficulties with daily routine
Mild difficulties with daily routine
No difficulties with daily routine
1.1b (if no COVID) Have you experienced any illness since the start of the pandemic?

Yes (skip to 1.4)
No (skip to 1.2)
1.1c (if had COVID twice) How long was it between your multiple bouts of COVID-19:

I had it twice within the same month
There was more than a month between my COVID-19 infections
1.1d (if had COVID twice) Did you have a positive COVID-19 test both times that you had COVID-19?

Yes
No
I was not tested
1.1e (if had COVID twice) Did you have a negative COVID-19 test in between your multiple bouts of COVID-19?

Yes
No
I was not tested
1.1f (if had COVID twice) Which of the following best describes your COVID-19 history:

The second time that I had COVID-19, the symptoms were much worse than the first time
The second time that I had COVID-19, the symptoms were more mild than the first time
The symptoms were of similar severity both times that I had COVID-19
Other (specify)
1.2 (if yes to COVID or any illness, including in vaccine questions above. If yes to COVID twice, repeat all parts of question 1.2 twice) Did you experience any of the following symptoms as part of your illness [check all that apply]: Series of Check boxes:

_Fever (temperature greater than 38 degrees Celsius or 100 degrees Fahrenheit)
_Cough with mucus / phlegm / sputum
_Dry cough (cough without mucus / phlegm / sputum)
_Sputum (phlegm) production
_Sore Throat
_Runny nose
_Red, sore or itchy eyes
_A lump in your throat
_Difficulty breathing or shortness of breath
_Severe difficulty breathing
_Pain with deep breaths
_Being woken up by shortness of breath
_Severe fatigue, such as inability to get out of bed
_Sore muscles
_Body aches
_Muscle weakness
_Bone or joint pain
_Lower back pain
_Nausea
_Vomiting
_Decrease in appetite
_Acid reflux
_Diarrhea
_Loss of sense of smell
_Loss of sense of taste
_Dizziness
_Difficulty balancing
_Headache
_Confusion
_Difficulty concentrating
_Decreased alertness
_Memory loss
_Insomnia (difficulty sleeping)
_Ringing in the ears (tinnitus)
_Pain in the chest or heart area
_Heart palpitations
_Noticing your heartbeat
_Intermittent unexplained elevated heart rate (tachycardia)
_Feeling very warm sometimes, then feeling very cold again
_A numb or tingling sensation somewhere in your body
_Feeling limp somewhere
_Feeling heavy in the arms or legs
_Rash
_Sensitive skin
_Blisters
_Discolored fingers or toes
_None of the above
_Other (please specify):
1.2.1 if any boxes checked for 1.2
What was your first symptom?
Answer: drop-box of same list as above
1.2.2 if any boxes checked for 1.2
Are you still suffering from any symptoms?
Answer: drop-box of same symptom list as above
1.2.3 if any boxes checked for 1.2
For approximately how many days did the symptom last?
1.2.4 if any boxes checked for 1.2
For approximately how many days, weeks or months were you or have you been so unwell that you stayed in bed or on the sofa (Fill in whichever number you remember best)?
Approximately how many days, weeks or months ago did you become ill? (Fill in whichever number you remember best)
1.3 Since being ill, have you been newly diagnosed with any of the following conditions or had any of the following procedures? [check all that apply]: Series of Check boxes:

_Blood clots
_Stroke or TIA
_Pulmonary embolism
_Heart attack (cardiac arrest)
_Heart failure
_Myocarditis
_Heart damage or scarring
_Pacemaker implant
_Lung fibrosis
_Lung damage or scarring
_Pneumonia
_Acute Respiratory Distress Syndrome (ARDS)
_Oxygen supplementation
_Dialysis
_Kidney damage
_Renal failure
_None of the above
_Other (please specify)
1.4 Have you been tested for COVID-19?

Answer
Yes, and it was positive (confirmed COVID-19)
Yes, and it was negative (did not confirm COVID-19)
Yes, and the result was inconclusive
Yes, and the result is still pending
No
Unsure
1.4a What kind of COVID-19 test did you have?

Antibody test (to tell if you’ve ever been infected)
Molecular or PCR test (to tell if you are currently infected)
Rapid or antigen test (to tell if you are currently infected, takes less than 1 hr)
Unsure
1.4b What type of sample was taken for your COVID-19 test?

Nasopharyngeal swab (long Q-tip deep into the nose) Anterior nares swab (short Q-tip shallowly into the nose)
Saliva (spit or sputum)
Blood
Other (please specify):
1.4c Why did you receive COVID-19 testing?

Symptoms
Exposure to infected person
Workplace or school-wide screening
Travel requirement
Part of medical procedure
Curiosity
Other (please specify)
1.4 Did you spend an overnight in the hospital or emergency room for coronavirus? Answer Yes/No
1.4.1 if yes to 1.4 Did you need oxygen (normally this would be administered through plastic tubes that are loosely inserted into your nose (i.e. nasal cannula), or through a mask applied over your face (BiPAP or CPAP)?

Answer Yes, oxygen via nasal cannula / Yes, BiPAP or CPAP mask / No
1.4.2 if yes to 1.4 Were you admitted to the Intensive Care Unit?

Answer Yes/No
1.4.3 if Yes to 1.4 Were you put on a mechanical ventilator?

Answer Yes/No
1.4.4 if Yes to 1.4 Were you diagnosed with pneumonia due to coronavirus?

Answer Yes No Unsure
1.4.5 if Yes to COVID

Did you have any of the following complications of coronavirus? Check all that apply.
Series of check boxes:
Septic shock
Multiple organ dysfunction/failure
Radiographic lung infiltrates (diagnosed via CXR or CT)
Hepatitis
Pancreatitis
Pleural effusion
Acute kidney failure
Kidney Insufficiency
Ascites / excess abdominal fluid
Encephalitis
Heart failure
Anemia
Vasculitis / blood vessel inflammation

Unsure
None of the above
Other (please specify)
1.4.6 if Yes to COVID Did a physician prescribe you a treatment for coronavirus? Check all that apply.

_Remdesivir
_Convalescent plasma
_Monoclonal antibodies
_Azithromycin
_Chloroquine
_Hydroxychloroquine
_Corticosteroids
_Lopinavir-Ritonavir
_Tocilizumab
_JAK inhibitor
_Unsure
_No
_Other (please specify)

We will now ask some questions about yourself to study whether you have been at increased or decreased risk of coronavirus due to the medications you take, any chronic conditions you may have, your daily behaviors, and other factors. All questions are voluntary.

2.1 What is your current weight (in pounds)? Answer: Number
2.2 What is your height in feet and inches? Answer: Feet and inches options list from 4’0” to 8’0”
2.3 Please check below if you’ve ever been diagnosed with any of the following conditions or had any of the following procedures [check all that apply]: Series of Check boxes:

_Cardiovascular (heart) disease
_High blood pressure / hypertension
_Asthma
_COPD (chronic obstructive pulmonary disease)
_Chronic bronchitis
_Cystic Fibrosis
_Other chronic lung disease
_Type 1 Diabetes
_Type 2 Diabetes
_Sleep apnea
_Use of a home CPAP (continuous positive airway pressure) device at home at night
_HIV
_Immunocompromised status
_Organ transplant
_Bone marrow transplant
_Rheumatoid arthritis
_Systemic lupus
_Erythematosus
_Multiple sclerosis
_Inflammatory bowel disease
_Celiac disease
_None of the above
_Other autoimmune or rheumatologic disease
Specify other automminue or rheumatologic disease If checked:
Please specify your other chronic lung disease
2.4 Do you regularly take any medications or supplements?

Yes / No
2.4a if Yes: Do you regularly take any of the following medications? Check all that apply.

Series of Check boxes:

_ACE-inhibitors (such as Lisinopril)
_Angiotension II Receptor Blockers (such as Losartan)
_Antibiotics
_Antiviral (such as the antimalarial hydroxychloroquine or HIV protease inhibitors (ritonavir, lopinavir, darunavir, atazanavir))
_Pollen allergy medications (such as Claritin, Zyrtec)
_Androgen deprivation therapy
_Asthma medication
_Light immunosuppressive medication (such as corticosteroids, DMARDs, anti-cytokine antibodies)
_Strong immunosuppressive medication (such as MMF, Tac, medications given to transplant patients)
_Medications called “biologics” or “monoclonal antibodies” such as abatacept (Orencia), adalimumab (Humira), anakinra (Kineret), certolizumab pegol (Cimzia), etanercept (Enbrel), golimumab (Simponi), infliximab (Inflectra, Remicade), rituximab (Rituxan), tocilizumab (Actemra), tofacitinib (Xeljanz), or upadacitinib (Rinvoq)
_Non-steroidal anti-inflammatory drugs (NSAIDs, such as Ibuprofen/Motrin, Naproxen/Aleve)
_Other pain/fever relievers (such as Aspirin, Paracetamol/acetaminophen)
_Blood thinners (such as Eliquis, Xarelto, Pradaxa)
_Vitamin D
_Bile acids (UDCA)
_None of the above
3.1 Have you smoked at least 100 cigarettes in your entire life? (There are 20 cigarettes in a pack)?

Answer Yes, I currently smoke / Yes, I used to smoke / No
3.1.1 if “Yes, I currently smoke” to 3.1:
When you smoke, how many cigarettes do you usually smoke daily?
Answer:
I don’t smoke regularly
1-10
11-20
21-30
31 or more
3.1.2 if “Yes, I used to smoke” to 3.1:
When you smoked, how many cigarettes did you usually smoke daily? Answer:
I don’t didn’t smoke regularly 1-10
11-20
21-30
31 or more
4 What is your blood type?

Answer A / B / AB / O / Don’t know
5 What is your Rh factor (i.e., + if your blood type is A+, - if your blood type is A-)

Answer + / - / Don’t know
6 What is your current age?

Answer Number
7.1 In what state do you live?

Drop-down list of states
7.1.1 What is your postcode/zip code?
Answer: free text
7.2 How many people do you live with at the moment? Do not count yourself. Answer: Number
7.3 How many children do you live with at the moment? Answer: Number
7.3a How many children do you live with who are attending in-person school or daycare during the pandemic? Answer: Number
7.4 Does anyone you share a home with, including yourself, currently work outside the home? Answer Yes / No
7.5 Did you get a flu vaccination (flu shot) in the past year? Answer: Yes / No

7.5a Did you experience any of the following symptoms after receiving the flu shot [check all that apply]:

Swelling at the injection site
Redness at the injection site
Severe pain at the injection site
Moderate pain at the injection site Muscle pain
Joint pain
Fatigue
Fever
Headache
Confusion or difficulty concentrating
Chills
Nausea
Vomiting
Intense hunger
Swollen lymph nodes / glands
Feeling unwell
Difficulty breathing
Swelling of your face or throat
A fast heartbeat
A bad rash all over your body
Dizziness
Difficulty sleeping
Delayed menstrual cycle
Early menstrual cycle
Other (please specify)
None
7.5b When at their worst, how much did the symptoms that you experienced after the flu shot interfere with your daily routine? Extreme difficulties / unable to perform daily routine Severe difficulties with daily routine Moderate difficulties with daily routine Mild difficulties with daily routine No difficulties with daily routine
7c How long after receiving the flu shot did these symptoms begin? Within 4 hours Within 3 days After more than 3 days
7d Once they started, how long did it take for the MOST SEVERE symptoms of the flu shot to go away?

Less than two days
Two to seven days
More than a week
7e Once they started, how long did it take for ALL of the symptoms of the flu shot to go away?

Less than two days
Two to seven days
More than a week
7.5f Have you ever experienced an adverse reaction to a vaccination?

No
Yes (please specify):
7.5g Are you allergic to any of the following?

Pollen
Pet / fur / feathers
Mold
Latex
Bee / wasp / insect
Dairy
Egg
Peanut
Tree Nut
Soy
Wheat/gluten
Fish
Shellfish
Medication
Other (please specify)
7.6 Have you been pregnant at any time since January 2020? Answer Yes / No
7.6.1 if Yes to 7.6 (if had COVID) Were you pregnant when you had COVID-19 (if had vaccine) Were you pregnant when you received the COVID-19 vaccine?
7.8 How many times in your life have you been diagnosed with pneumonia?

Answer Number
7.9 How many times in your life have you been diagnosed with bronchitis?

Answer Number
7.10 In general, how many times per year do you have a cold?

Answer Number
7.11 In general, how many times per year do you have sinusitis (sinus infections)?

Answer Number
7.12 In general, how many times per year do you have other upper respiratory tract infections?

Answer Number
7.13 Are you frequently around young children (for example, do you have young children or are you an elementary school teacher)? Check all that apply. Answer Yes, I have young children / Yes, I teach young children / Yes, other / No
7.14 How many hours of sleep do you usually get per night? Answer: Number
7.15 What is your highest level of educational attainment? Answer: Doctoral degree / Master’s Degree / Bachelor’s Degree (4 yrs) / Associate’s Degree (2 yrs) / Some college, no degree / High school diploma / Some high school, no diploma / Elementary/primary school
7.16 Have you ever had a tuberculosis (BCG) vaccine? Answer: Yes / No / Unsure
8.1 Has a close blood relative of yours (parent, child or sibling) been diagnosed with COVID-19?

Answer: Yes, my parent | Yes, my sibling | Yes, my child | No
8.1.1 if Yes to 8.1
Were you in physical contact with this relative within two weeks of their illness? Answer Yes No
8.1.2 if yes to 8.1
How severe was the illness of your close blood relative with coronavirus? If multiple close blood relatives had the virus, then please check as many boxes as apply. Series of check boxes:

_Mild case (no medical action taken)
_Contacted or saw physician
_Admitted to hospital
_Spent overnight in hospital
_Admitted to Intensive Care Unit
_Underwent mechanical ventilation
_Passed away from coronavirus
_Unsure
8.10 Are you a healthcare worker who comes into contact with patients?
Yes, and I work with coronavirus patients
Yes, but I do not come into contact with coronavirus patients
No
8.10 Are you an essential worker who physically goes into work during the pandemic?
Answer: Yes/No
8.10 Do you currently take precautions when you go out in public? Check all that apply.
Series of check boxes:
Wear face mask
Keep 6 feet from others
Avoid indoor public spaces
Avoid indoor restaurants and bars
Use hand sanitizer
Perform frequent hand washing
Avoid touching face
None of the above
9. Please check below if you’ve ever had any of the following health conditions, procedures or treatments [check all that apply]: Series of Check boxes:

_Liver disease
_Kidney disease or renal insufficiency
_Chronic muscle disease
_Depression
_Anxiety disorder
_Other mental health condition, such as schizophrenia
_Dementia
_Parkinson’s Disease
_Alzheimer’s Disease
_Other Neurological disease
_Balloon angioplasty or percutaneous coronary intervention
_Coronary artery bypass
_Congestive heart failure
_Myocardial infarction
_Peripheral vascular disease
_Stroke
_Arrythmia
_Hepatitis
_Pancreatitis
_Pleural effusion
_Ascites / excess abdominal fluid
_Leukemia
_Breast cancer
_Prostate cancer
_Bladder cancer
_Colon and rectal cancer
_Endometrial cancer
_Kidney cancer
_Liver cancer
_Melanoma
_Non-Hodgkin Lymphoma
_Pancreatic Cancer
_Thyroid Cancer
_Other cancer
_Other chronic disease
_None of the above

9.1 If “Other chronic disease” is checked, please specify
Answer: free text
9.2 If “Other cancer” is checked, please specify
Answer: free text
9.3 If “Other mental health condition” is checked, please specify
Answer: free text
9.4 If “Other neurological condition” is checked, please specify
Answer: free text
10. How often do you consume alcoholic beverages?

Answer:
Never
Once a month or less
2-4 times a month
2-3 times per week
4 times or more per week
11. What is your total annual household income? Under $15,000 Between $15,000 and $29,999 Between $30,000 and $49,999 Between $50,000 and $74,999 Between $75,000 and $99,999 Between $100,000 and $149,000 Between $150,000 and $199,999 Over $200,000 Prefer not to answer

Thank you! We will send you additional surveys in the future that may ask you to update some of these answers as time passes. While we don’t want to take too much of your time, you may hear from us every month or so while we continue to study this incredibly important matter.

**Subsequent surveys given at a later date use the same questions from the first survey but remove ones that are not expected to change much over time, such as medical history and age.**

## References

1. Chapin-Bardales, J., Gee, J. & Myers, T. Reactogenicity Following Receipt of mRNA-Based COVID-19 Vaccines. JAMA 325, 2201–2202 (2021).

2. Polack, F. P. et al. Safety and Efficacy of the BNT162b2 mRNA Covid-19 Vaccine. N. Engl. J. Med. 383, 2603–2615 (2020).

3. Baden, L. R. et al. Efficacy and Safety of the mRNA-1273 SARS-CoV-2 Vaccine. N. Engl. J. Med. 384, 403–416 (2021).

4. Menni, C. et al. Vaccine side-effects and SARS-CoV-2 infection after vaccination in users of the COVID Symptom Study app in the UK: a prospective observational study. Lancet Infect. Dis. 21, 939–949 (2021).

5. COVID-19 Host Genetics Initiative. Mapping the human genetic architecture of COVID-19. Nature (2021) doi:10.1038/s41586-021-03767-x.

6. Casanova, J.-L., Su, H. C. & COVID Human Genetic Effort. A Global Effort to Define the Human Genetics of Protective Immunity to SARS-CoV-2 Infection. Cell 181, 1194–1199 (2020).

7. Helix’s Exome+ Performance White Paper. (2019).

8. Introducing the Helix DNA Discovery Project. https://blog.helix.com/helix-dna-discovery-project/ (2018).

9. Grzymski, J. J. et al. Population Health Genetic Screening for Tier 1 Inherited Diseases in Northern Nevada: 90% of At-Risk Carriers are Missed. bioRxiv 650549 (2019) doi:10.1101/650549.

10. Cirulli, E. T. et al. Genome-wide rare variant analysis for thousands of phenotypes in over 70,000 exomes from two cohorts. Nat. Commun. 11, 542 (2020).

11. Zheng, X. et al. HIBAG--HLA genotype imputation with attribute bagging. Pharmacogenomics J. 14, 192–200 (2014).

12. Mbatchou, J. et al. Computationally efficient whole genome regression for quantitative and binary traits. Cold Spring Harbor Laboratory 2020.06.19.162354 (2020) doi:10.1101/2020.06.19.162354.

13. Sadoff, J. et al. Safety and Efficacy of Single-Dose Ad26.COV2.S Vaccine against Covid-19. N. Engl. J. Med. 384, 2187–2201 (2021).

14. Gonzalez-Galarza, F. F. et al. Allele frequency net database (AFND) 2020 update: gold-standard data classification, open access genotype data and new query tools. Nucleic Acids Res. 48, D783–D788 (2020).

15. McInnes, G. et al. Global Biobank Engine: enabling genotype-phenotype browsing for biobank summary statistics. Bioinformatics 35, 2495–2497 (2019).

16. Pacho, A. et al. HLA haplotypes associated with hemochromatosis mutations in the Spanish population. BMC Med. Genet. 5, 25 (2004).

17. Mallal, S. et al. Association between presence of HLA-B*5701, HLA-DR7, and HLA-DQ3 and hypersensitivity to HIV-1 reverse-transcriptase inhibitor abacavir. Lancet 359, 727–732 (2002).

18. Illing, P. T. et al. Immune self-reactivity triggered by drug-modified HLA-peptide repertoire. Nature 486, 554–558 (2012).

19. Chung, S. et al. GWAS identifying HLA-DPB1 gene variants associated with responsiveness to hepatitis B virus vaccination in Koreans: Independent association of HLA-DPB1*04:02 possessing rs1042169 G - rs9277355 C - rs9277356 A. J. Viral Hepat. 26, 1318–1329 (2019).

20. Png, E. et al. A genome-wide association study of hepatitis B vaccine response in an Indonesian population reveals multiple independent risk variants in the HLA region. Hum. Mol. Genet. 20, 3893–3898 (2011).

21. Pöyhönen, L., Bustamante, J., Casanova, J.-L., Jouanguy, E. & Zhang, Q. Life-Threatening Infections Due to Live-Attenuated Vaccines: Early Manifestations of Inborn Errors of Immunity. J. Clin. Immunol. 39, 376–390 (2019).

22. Lichtenauer-Kaligis, E. G. R. et al. Severe Mycobacterium bovis BCG infections in a large series of novel IL–12 receptor β1 deficient patients and evidence for the existence of partial IL–12 receptor β1 deficiency. European Journal of Immunology vol. 33 59–69 (2003).

23. Tartof, S. Y. et al. Effectiveness of mRNA BNT162b2 COVID-19 vaccine up to 6 months in a large integrated health system in the USA: a retrospective cohort study. Lancet 398, 1407–1416 (2021).

